# Evaluation and classification of severity for 176 genes on an expanded carrier screening panel

**DOI:** 10.1101/2019.12.14.19014951

**Authors:** Aishwarya Arjunan, Holly Bellerose, Raul Torres, Rotem Ben-Shachar, Jodi D. Hoffman, Brad Angle, Robert Nathan Slotnick, Brittany N. Simpson, Andrea M. Lewis, Pilar L. Magoulas, Kelly Bontempo, Jeanine Schulze, Jennifer Tarpinian, Jessica A Bucher, Richard Dineen, Allison Goetsch, Gabriel A. Lazarin, Katherine Johansen Taber

## Abstract

**Background:** Severity is an important factor for inclusion of diseases on expanded carrier screening (ECS) panels. Here, we applied a validated algorithm that objectively classifies diseases into severity categories to 176 genes on a clinically available ECS panel. We then mapped disease traits from the algorithm to severity-related criteria cited by the American College of Obstetricians and Gynecologists (ACOG).

**Methods:** Eight genetic counselors (GCs), followed by four medical geneticists (MDs), applied the algorithm to subsets of the 176 genes. MDs and GCs then determined which disease traits met ACOG severity criteria.

**Results:** Upon initial GC and MD review, 107/176 genes (61%) and 133/176 genes (76%), respectively, had concordant classifications, with consensus reached for all genes. Final severity classifications were 68 (39%) profound, 71 (40%) severe, 36 (20%) moderate, and one (1%) mild. The vast majority of genes (170 out of 176) met at least one of ACOG’s four severity criteria.

**Conclusion:** This study classified the severity of a large set of Mendelian genes by collaborative clinical expert application of an algorithm. Further, it clarified and operationalized difficult to interpret ACOG severity criteria via mapping of disease traits, thereby promoting consistency of ACOG criteria interpretation across laboratories.

**What’s already known about this topic?:**

- Disease severity is an important consideration for disease inclusion on expanded carrier screening panels.
- An algorithm that objectively classifies diseases into severity categories has been published and validated.

**What does this study add?:**

- 176 genes were classified into severity categories.
- The algorithm was used to bring clarity to American College of Obstetricians and Gynecologist’s (ACOG’s) severity criteria that are not easily interpretable.
- 170 of 176 genes met at least one of ACOG’s severity criteria.

**Data Availability Statement:** The data that support the findings of this study have been completely reported in this manuscript and shared in the Figures and Supplementary Material.

## Introduction

Carrier screening identifies couples at risk of having offspring affected by a genetic disease, thereby informing reproductive decision-making and pregnancy management. Expanded carrier screening (ECS) accomplishes this for dozens to hundreds of diseases, in comparison to traditional ethnicity-based screening approaches meant to detect a limited number of conditions prevalent within those ethnic groups (1). The American College of Obstetricians and Gynecologists (ACOG) considers ECS to be an acceptable screening strategy and has proposed that conditions selected for screening panels should meet several of seven criteria for disease inclusion on ECS panels (2). Four of these criteria address disease severity, stating that conditions should: 1. have a detrimental effect on quality of life, 2. cause cognitive or physical impairment, 3. have an onset early in life, or 4. require surgical or medical intervention (2). Similarly, the American College of Medical Genetics and Genomics (ACMG) position statement on ECS states that “disorders should be of a nature that most at-risk patients and their partners identified in the screening program would consider having prenatal diagnosis to facilitate making decisions around reproduction” (3). However, individuals have varying perceptions of the concept of severity based on their valuation of phenotypic traits (4, 5).

Disease severity inclusion criteria for ECS panels have not been widely studied. However, multiple studies have shown that disease categorization is helpful for patients in understanding the types of diseases included on ECS panels, and facilitates reproductive decision-making (5-7). In 2014, Lazarin et al. published an algorithm to objectively categorize disease severity (4). In the study, 192 health care providers rated 13 individual disease traits related to disease severity, independent of any named genetic disease. Disease traits (e.g., shortened lifespan, intellectual disability), as opposed to diseases themselves, were evaluated. This is because, despite health care providers’ potential lack of familiarity with many rare genetic diseases, the traits associated with them are often encountered by health care providers regardless of the etiology. The analysis established and validated an algorithm that can be applied to a disease, with its given set of individual phenotypic traits, resulting in a classification of the condition into one of four severity categories: profound, severe, moderate, and mild (4).

To date, no published studies have rigorously applied the algorithm using health care providers with specialized knowledge of genetic disease, nor has it been applied to the large number of diseases currently included on many ECS panels. In this study, we aimed to assign an objective severity classification to 176 genetic diseases in order to inform consideration of their inclusion on ECS panels. We engaged board-certified pediatric genetic counselors and medical geneticists in clinical practice who have experience treating patients affected with conditions commonly found on ECS panels. Furthermore, we mapped disease traits defining the algorithm’s severity categories to ACOG’s four severity criteria in order to provide objective definitions of the criteria.

## Materials and Methods

### Institutional Review Board Considerations

This study did not use patient samples or results and therefore was not subject to review by an institutional review board or ethics committee.

### Gene Classifications

Eight pediatric genetic counselors (GCs) followed by four medical geneticists (MDs) applied a validated severity algorithm (4) to randomly assigned subsets of 176 genes offered on a commercially available ECS panel (Foresight, Myriad Women’s Health). The algorithm was applied to *genes*, rather than *diseases*, because some conditions are caused by multiple genes (e.g. Usher Syndrome, Fanconi Anemia, Niemann Pick Disease) and some genes are associated with multiple conditions (e.g. *HBB, FKTN, MYO7A*). The algorithm (described in Figure 1) was applied in a two-step review and consensus process as described in Figure 2 and Supplementary Text S1. Honoraria were provided to each GC and MD participant. Permutation testing was conducted to determine whether particular individuals within each GC (1-4) or MD (1-2) pair consistently classified disease genes as higher or lower in severity than their counterpart (see Supplementary Text S2).

**Figure 1.**
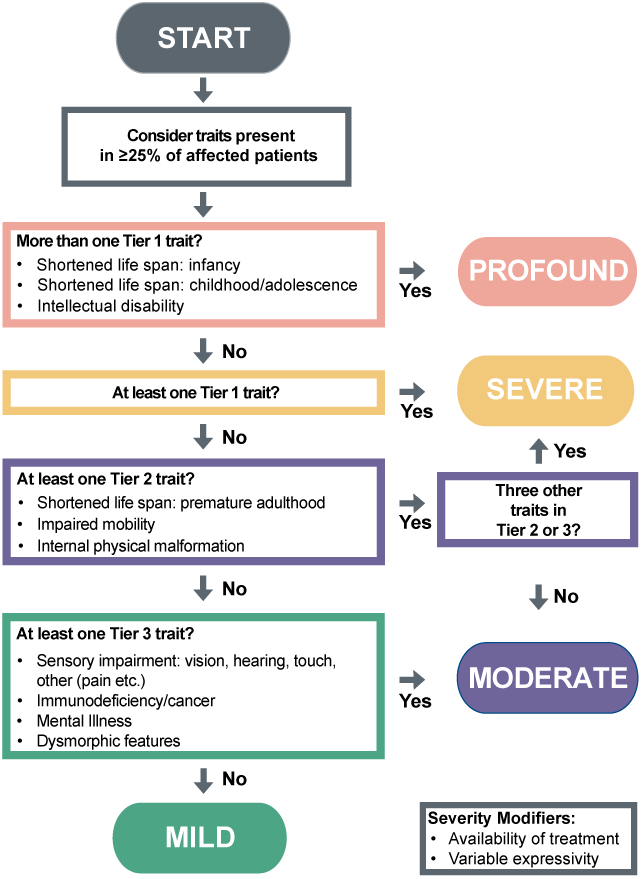
Severity classification algorithm evaluating specified disease traits (published by (4)). Each GC and MD used the algorithm to independently review and classify genes and their associated phenotype arising in at least 25% of individuals in the untreated state, as profound, severe, moderate, or mild.

**Figure 2.**
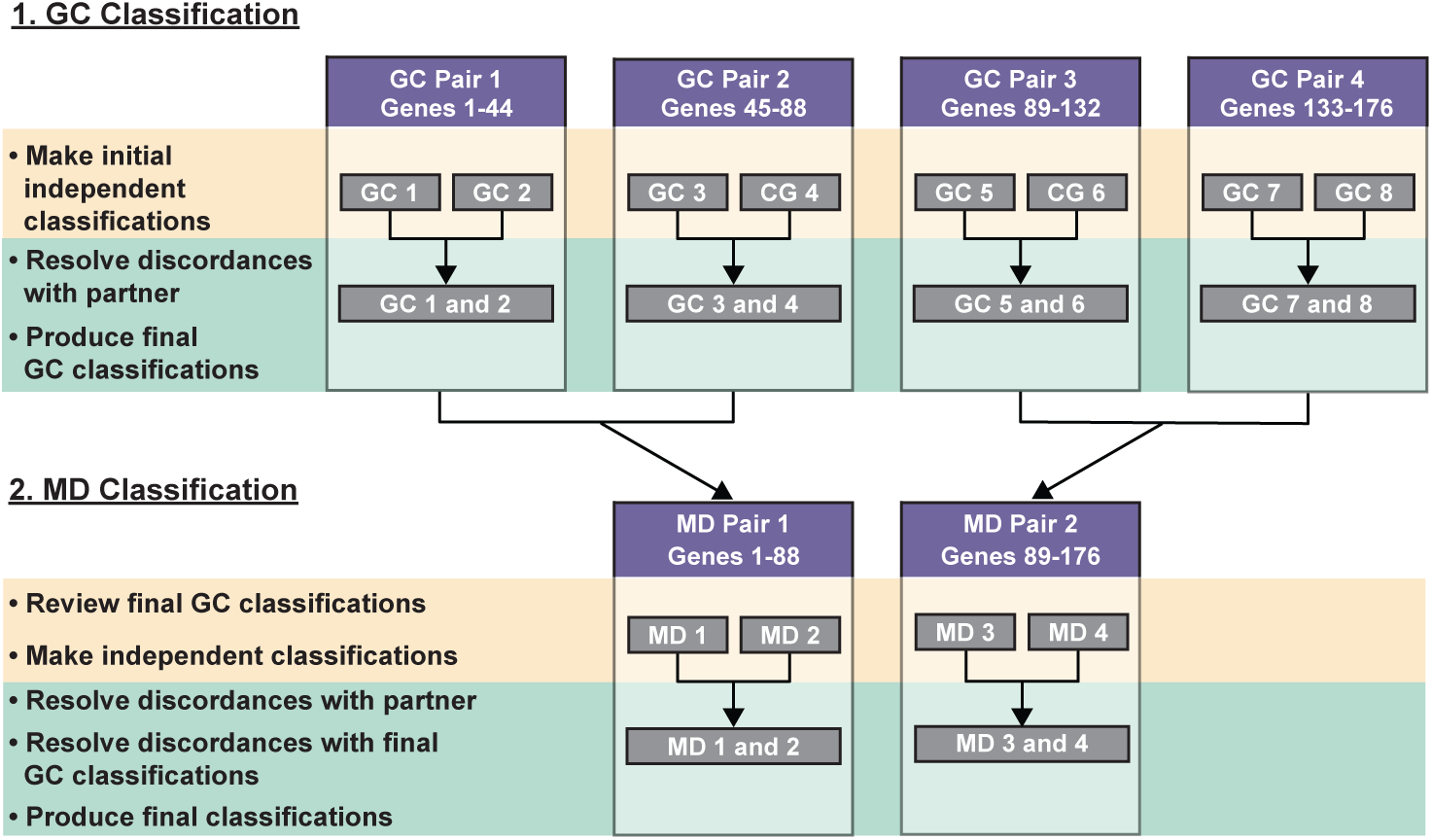
Workflow of the severity classification process. Eight genetic counselors (GCs) were divided into four pairs; each pair was asked to review 44 genes. Once the GC pairs finished their initial review and resolved discordances, the classification process was then passed to the four medical geneticists (MDs) for review and final classification. Gene numbers in this figure correspond to the gene numbers in Table 1.

### Mapping Disease Traits to ACOG criteria

Following GC and MD gene classifications, each disease trait assessed in the algorithm (see Figure 1) was mapped collaboratively by the MDs and GCs to the four ACOG severity criteria: 1. a detrimental effect on quality of life, 2. cognitive or physical impairment, 3. onset early in life, and 4. requires surgical or medical intervention. Disease traits were mapped to ACOG severity criteria only if all GCs and MDs assessing each gene agreed on the traits. To assess ACOG’s “detrimental effect on quality of life” criterion, we considered “quality of life” to be defined by the domains included in the Pediatric Quality of Life Inventory (PedsQL) 4.0 Generic Core Scales (physical functioning, emotional functioning, social functioning, and school functioning) (8) and PedsQL Infant Scales (physical functioning, physical symptoms, emotional functioning, social functioning, and cognitive functioning) (9). Disease traits that negatively impact these PedsQL domains were then mapped to the “detrimental effect on quality of life” criterion. We considered ACOG’s “cause cognitive or physical impairment” criterion to be captured by the PedsQL Generic Core and Infant Scales domains of physical functioning and physical symptoms (physical impairment) and school functioning and cognitive functioning (cognitive impairment) (8, 9). To interpret ACOG’s “onset early in life” criterion, we used the American Academy of Pediatrics’ (AAP) definitions of infancy (birth to age 2 years of age), childhood (2-12 years of age), and adolescence (12-21 years of age) (10). Though the severity algorithm uses shortened lifespan as a disease trait rather than age of onset, we reasoned that a condition that results in death before the end of adolescence (age 21 years) would be considered “early onset.” Thus, to simplify the mapping, tier 1 traits of “shortened life span: infancy” and “shortened life span: childhood/adolescence” were combined into one trait. We interpreted “require surgical or medical intervention” as only interventions that are delivered within the context of the health care setting and that are required to treat, delay, halt the onset of, or lessen the severity of disease symptoms.

**Table 1.**
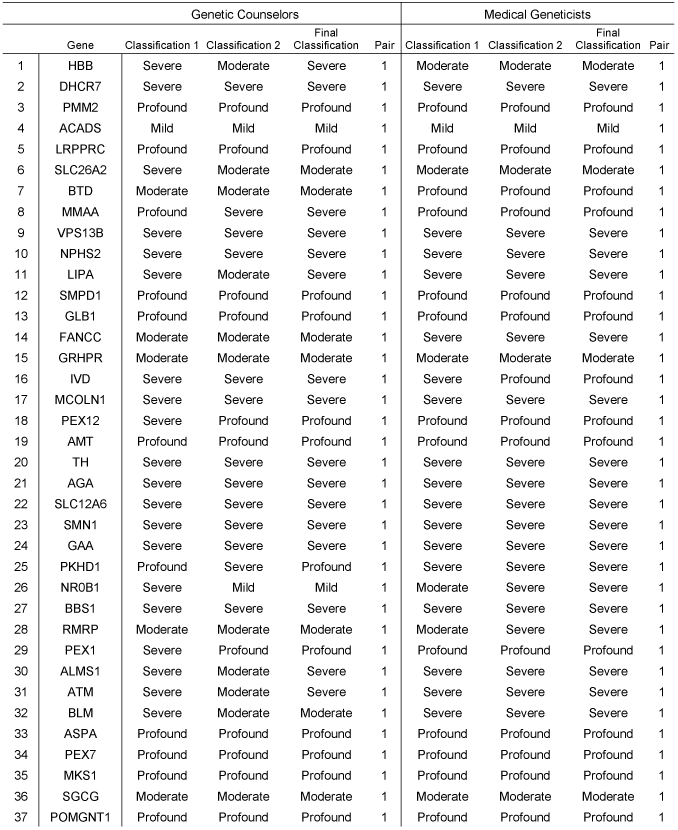
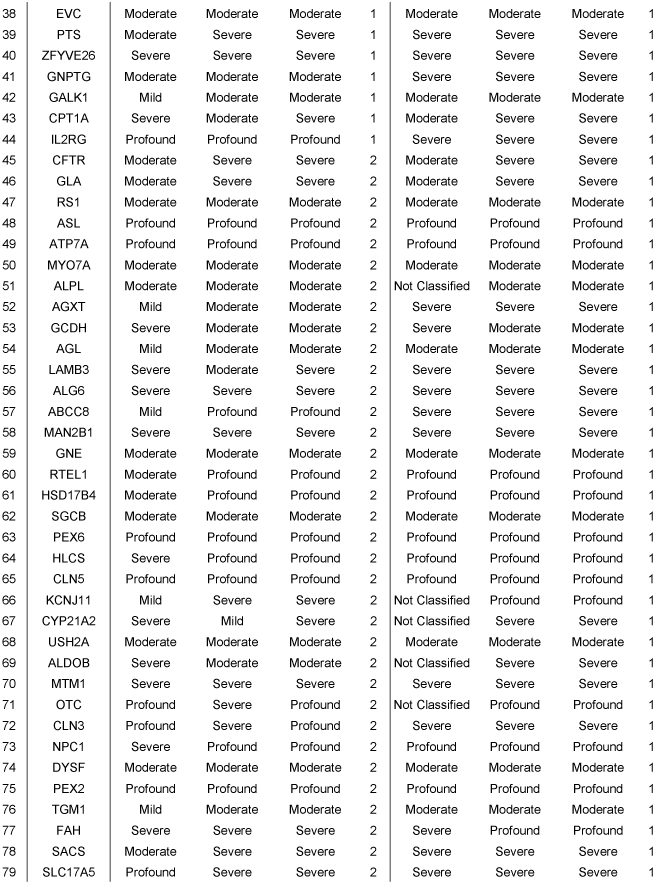
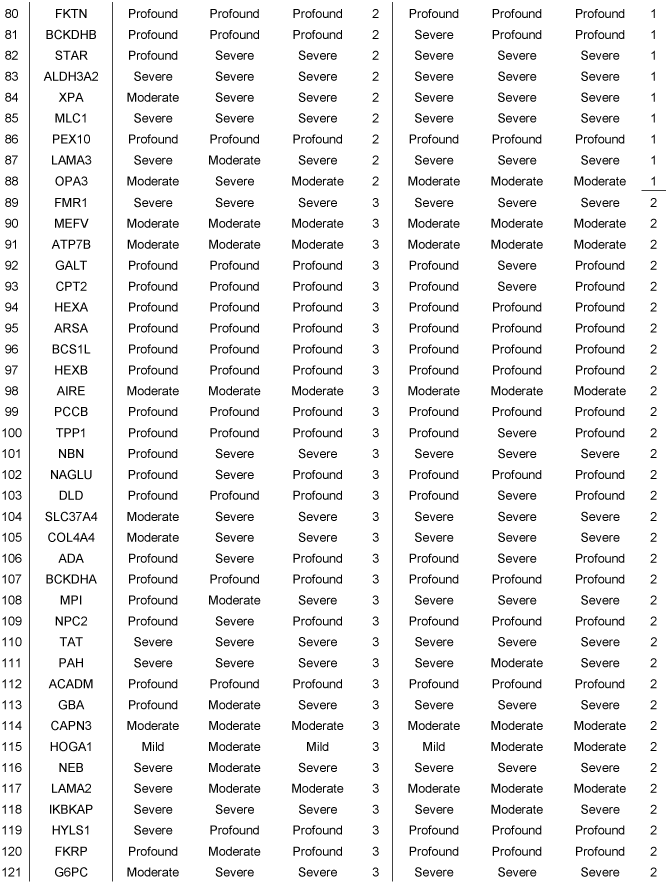
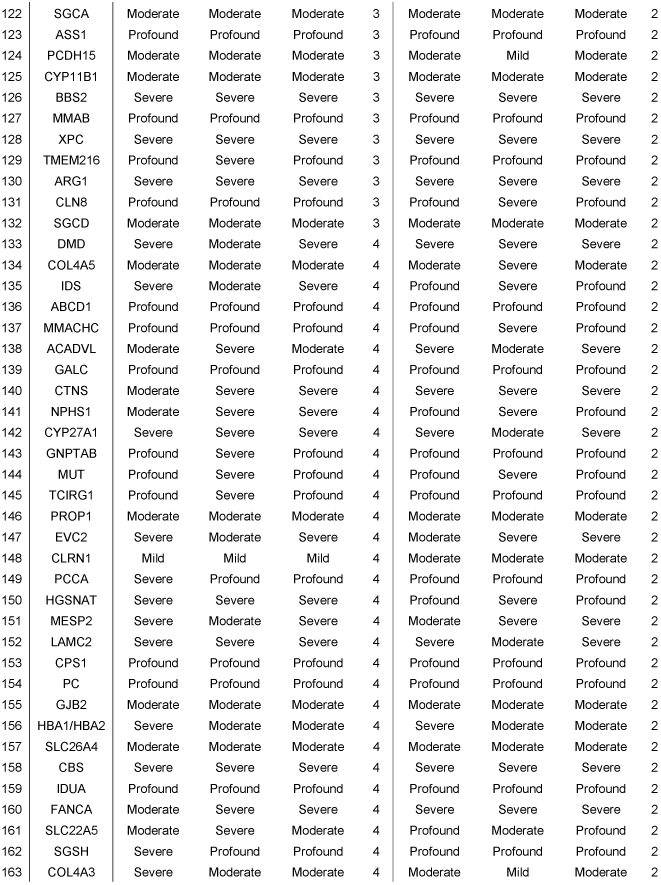
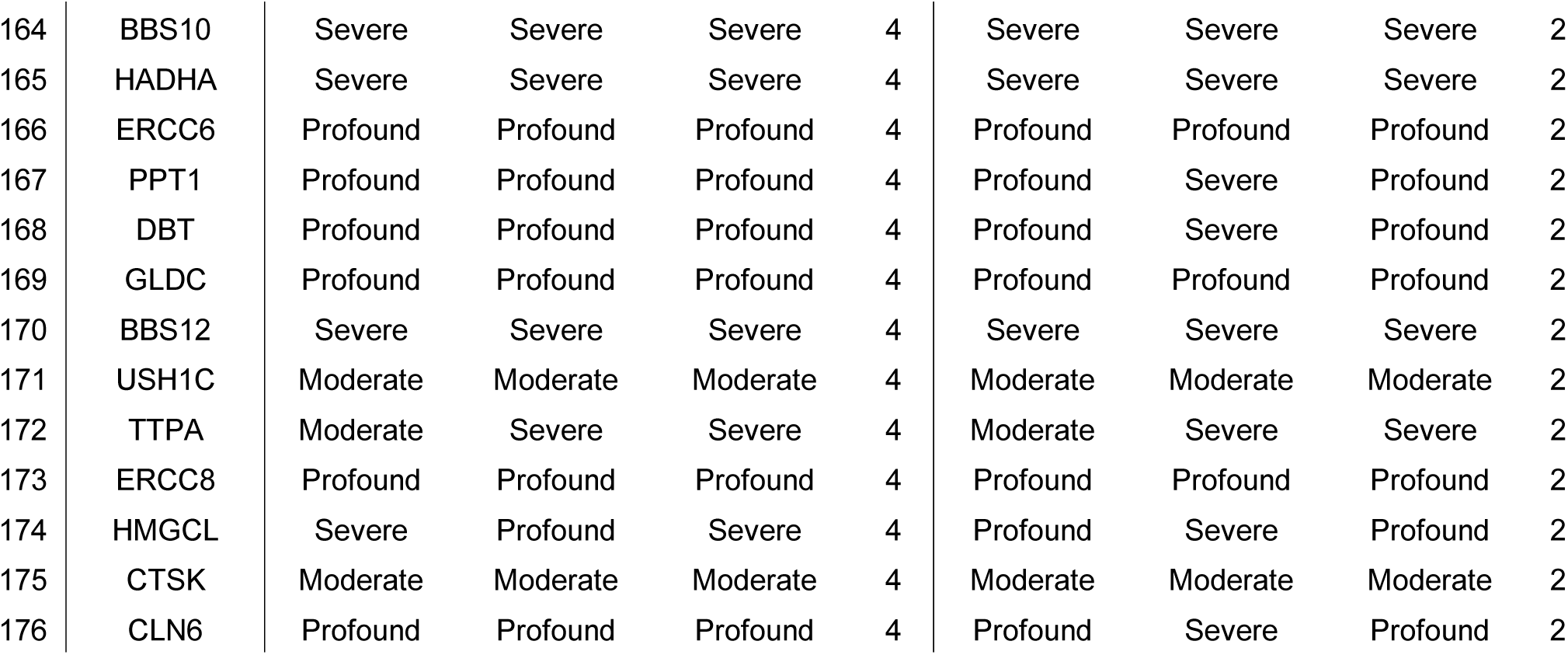
Initial and final severity classifications between pairs of genetic counselors (GCs) and medical geneticists (MDs).

### Data Analysis

Descriptive statistics were used to characterize general data trends. Statistical significance between proportions was determined using chi-squared analysis; a result was considered significant when p□<□0.05.

## Results

### Genetic Counselor Review and Classification

After initial review across all GC pairs, 107 of the 176 disease-associated genes (60.8%) had concordant severity classifications (Figure 3A, Table 1). Within the four GC pairs, concordances were 68.2% (30/44 genes), 47.7% (21/44 genes), 65.9% (29/44 genes), and 61.4% (27/44 genes), respectively (Figure 3A, Table 1). With the exception of four genes (*NR0B1, ABCC8, KCNJ11, CYP21A2*), all discordant classifications were within one level of classification (Figure 3A, Table 1). A permutation test was conducted to determine whether any individual within each GC pair tended to classify genes as more or less severe than their counterpart and found no significant difference (all p > 0.05; Supplementary Figure 1). Reasons for discordances noted by the GC pairs included difficulties in determining the primary phenotype associated with a specific disease with varying severity, difficulty discerning the level of intellectual disability associated with the disease from the available literature, and inability to locate published data regarding life expectancies and phenotypic differences in various forms of the conditions. After final review of discordances and consensus on final classifications between each GC pair, 65 genes (36.9%) were categorized as profound, 65 (36.9%) as severe, 42 (23.9%) as moderate, and four (2.3%) as mild (Table 1).

**Figure 3.**
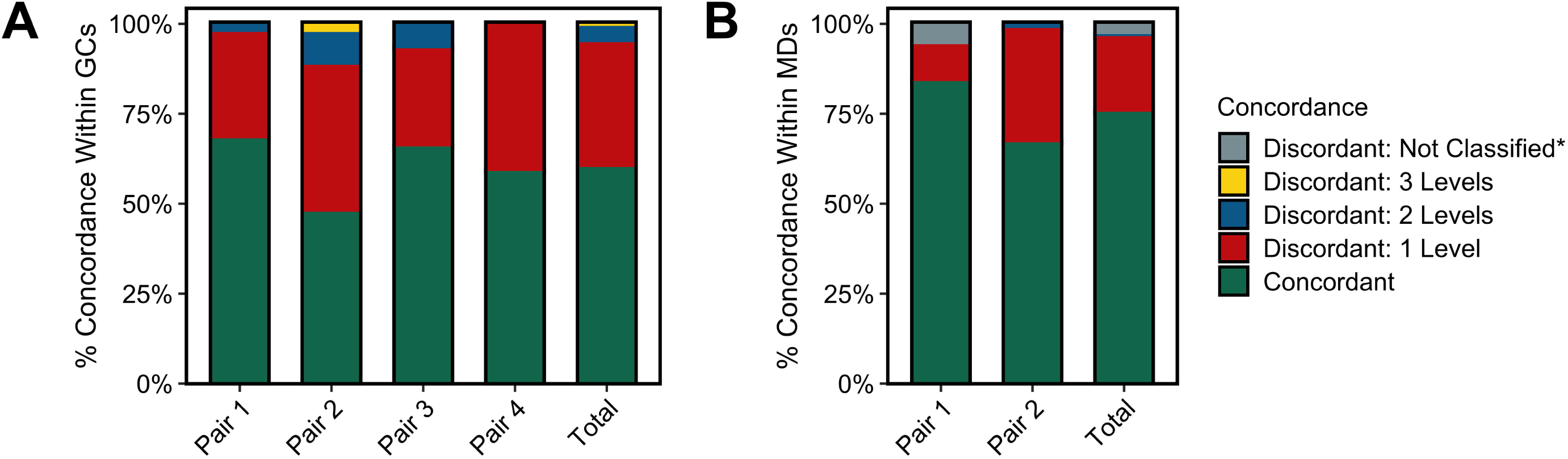
Concordance after initial gene severity classification. Concordance of initial severity classifications within each GC pair and in total among all GCs (A), and concordance of initial severity classification within each MD pair and in total among all MDs (B). Levels of discordance are indicated by color (pink: concordant, green: discordant by one level, blue: discordant by two levels, purple: not classified).

### Medical Geneticist Review and Classification

After initial review across both MD pairs, 133 of the 176 disease-associated genes (76%) had concordant classifications (Figure 3B, Table 1). Within each MD pair, concordances were 84.1% (74/88 genes, MD Pair 1) and 67% (59/88 genes, MD Pair 2) (Figure 3B, Table 1). One member of MD Pair 1 did not definitively classify five genes; these were considered discordances. All but one discordant classification (*SLC22A5*) were within one level of classification (Figure 3B, Table 1). A permutation test was conducted (similar to that for GCs) to determine whether any individual within each MD pair tended to classify genes as more or less severe than their counterpart and found no significant difference (all p > 0.05; Supplementary Figure 1). Reasons for discordances included no definitive classification, multiple phenotypes associated with the gene, unknown percentages of individuals with intellectual disability or shortened lifespan, and difficulties in determining life expectancy in the untreated state for conditions for which treatment is available. After final MD review of discordances and consensus on final classifications, 68 (38.6%) genes were categorized as profound, 71 (40.3%) as severe, 36 (20.5%) as moderate, and one (0.6%) as mild (Table 1). Comparison of GC and MD classifications revealed no significant differences in the number of genes ultimately classified within each severity category (36.9% vs. 38.6% profound, p=0.74; 39.6% vs. 40.3% severe, p=0.51; 23.9% vs. 20.5% moderate, p=0.44; 2.3% vs. 0.6% mild, p=0.18, chi-squared test).

### Disease Traits and Their Relationships to ACOG Severity Criteria

Disease traits most frequently cited by GCs and MDs across all genes as they applied the severity algorithm were sensory impairment (85 genes), intellectual disability (70 genes), and impaired mobility (67 genes) (Figure 4A). The disease traits that were least frequently cited were immunodeficiency/cancer (12 genes) and mental illness (9 genes) (Figure 4A). The list of genes and their assigned disease traits are provided in Supplementary Table 1.

**Figure 4.**
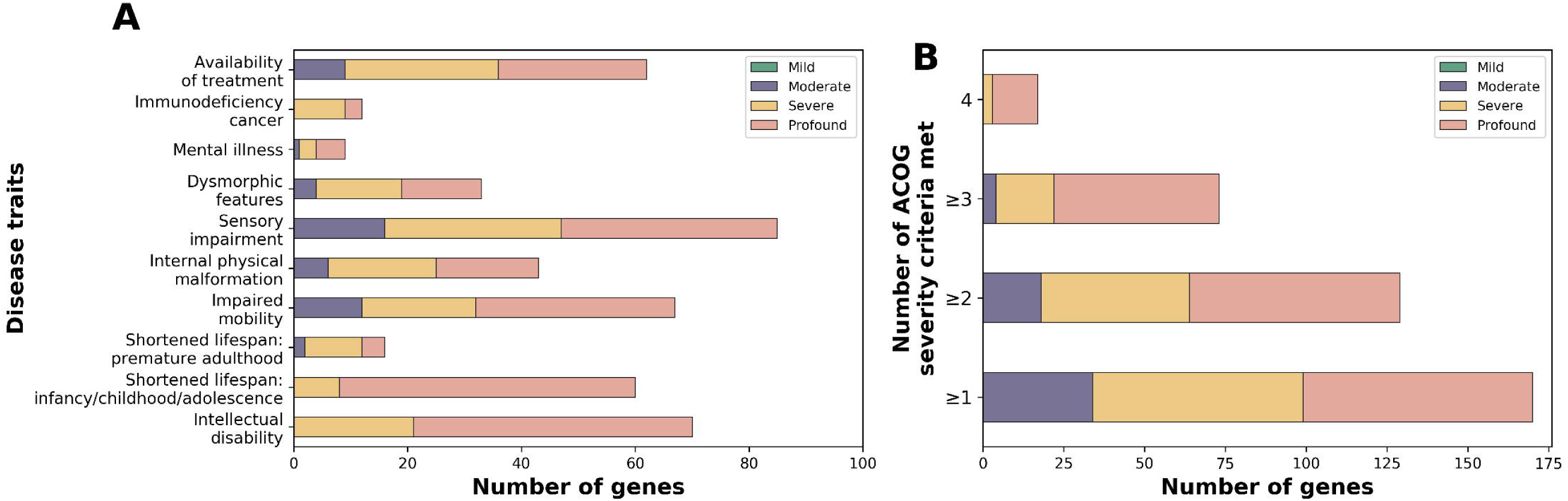
Genes classified by disease trait and ACOG severity criteria. (A) Genes classified by disease trait. (B) Genes classified by number of ACOG severity criteria met.

To determine which of the diseases evaluated in this study met ACOG’s four severity-related criteria, MDs and GCs mapped the disease traits from the severity algorithm to the ACOG criteria using validated scales and definitions (See Methods). The disease traits and their mapping to each ACOG severity criteria are described in Table 2. The list of genes and their ACOG severity criteria classification are provided in Supplementary Table 1.

**Table 2.** Mapping of the algorithm’s disease traits to ACOG severity criteria.

| ACOG Severity Criteria | Have a detrimental effect on quality of life | Cause cognitive or physical impairment | Have an onset early in life | Require surgical or medical intervention |
| --- | --- | --- | --- | --- |
| **Algorithm Disease Traits** | Intellectual disability<br>Impaired mobility<br>Internal physical malformation<br>Sensory impairment<br>Dysmorphic features<br>Mental illness<br>Immunodeficiency/<br>cancer | Intellectual disability<br>Impaired mobility | Shortened lifespan:<br>infancy/<br>childhood/adolescence | Availability of treatment |

The “detrimental effect on quality of life” criterion was the most frequently met ACOG criterion (155 [88.1%] genes; 65 profound, 57 severe, and 33 moderate, Supplementary Figure 2).This was followed by the “cause cognitive or physical impairment” criterion (97 [55.1%] genes; 54 profound, 31 severe, and 12 moderate, Supplementary Figure 2). Seventy-five (42.6%) genes met the “onset early in life” criterion (56 profound, 17 severe, and 2 moderate, Supplementary Figure 2). The least frequently met ACOG criterion was “require surgical or medical intervention” (62 [35.2%] genes; 26 profound, 27 severe, and 9 moderate, Supplementary Figure 2). The number of ACOG criteria met across all conditions was proportional to the categorized severity of the gene: on average, profound genes met 2.8 ACOG severity criteria, severe genes met 1.9 criteria, moderate conditions met 1.6 traits, and the one mild condition met 0 criteria.

In total, 170 out of 176 genes (96.6%) met at least one of the four ACOG severity criteria, 129 genes (73.3%) met at least two ACOG severity criteria, 73 genes (41.5%) met at least 3 ACOG severity criteria, and 17 genes (9.7%) met all four criteria (Figure 4B). Cystic fibrosis and spinal muscular atrophy, two conditions recommended for panethnic screening by ACOG (11), met one and four criteria, respectively (Supplementary Table 2). Of the remaining six genes that did not meet at least one of the four ACOG severity criteria, one was classified as mild, two were classified as moderate, and three were classified as severe (Figure 4B, Supplementary Table 1). The results of this disease trait-ACOG criteria mapping show that 170 of the 175 (97.1%) genes that were classified as profound, severe, and moderate met at least one of the ACOG severity criteria (Figure 4B, Supplementary Table 1).

## Discussion

Here, we have demonstrated the application of a severity classification algorithm applied by health care providers with expertise in pediatric and genetic disease to systematically classify the severity of 176 genes on a commercially available ECS panel. In addition, heath care providers operationalized the ACOG severity criteria via mapping of disease traits to the criteria, thereby determining which of the genes analyzed in this study met the criteria.

### Bringing Clarity to Severity Criteria in Guidelines

Guidelines stipulate disease severity as an important factor in the selection of conditions for ECS panel inclusion yet describe severity in terms that are not easily interpreted. Our study brings more clarity to ACOG’s severity criteria; MDs and GCs who are experts in rare disease unanimously agreed on the mapping of disease traits to the criteria. When evaluating 176 genes disease traits, 170 met at least one of ACOG’s severity criteria (Figure 4B), suggesting that they are appropriate to consider for inclusion on ECS panels.

MDs and GCs mapping disease traits to ACOG’s four severity criteria interpreted the criteria based on definitions available in the literature, whenever possible, to avoid assumptions about ACOG’s intent. For example, ACOG’s “detrimental effect on quality of life” criterion was assessed using a definition of “quality of life” developed based on domains in the PedsQL, a validated instrument for assessing quality of life in infants and children with chronic and acute disease (8, 9). Social and family support, environment, and socioeconomic status may modify detriments to the quality of life experienced by those with genetic disease, but were not included in the evaluation of the ACOG criteria because they are not clearly accounted for by the PedsQL instrument. Interpretation of the “onset early in life” criterion was based on the age limit of adolescence as defined by the AAP (10). To unambiguously map “shortened lifespan” (the trait assessed in the algorithm) to age of onset, only conditions that result in death before the end of adolescence were considered to meet the criterion. Therefore, genetic conditions that result in death during adulthood but that have an onset during or before adolescence, such as familial dysautonomia and glycogen storage disease type 1a, did not meet the “onset early in life” criterion, even though ACOG implicitly acknowledges the severity of such conditions by suggesting both familial dysautonomia and glycogen storage disease type 1a as examples of conditions to include on ECS panels (11). “Require surgical or medical intervention” was defined as interventions delivered in the formal health care setting that are required to treat, delay, halt the onset of, or lessen the severity of disease symptoms. For example, surgery to correct a cardiac defect and treatment with chelating medications were considered to meet the ACOG criterion. Palliative or supportive care, as well as interventions delivered outside the context of the medical setting such as specialized early education for those with learning disabilities were not considered to meet the criterion, though their benefits have been demonstrated (12).

While the majority of genes met at least one ACOG criteria, narrow interpretation of ACOG criteria resulted in 8.5% of profound conditions and 28.0% of severe conditions, including cystic fibrosis and Bloom syndrome, meeting only one ACOG criterion. Cystic fibrosis met the “require surgical or medical intervention” criterion because treatments are available to lessen the severity of disease symptoms. However, it does not usually cause death before the end of adolescence (the trait corresponing to ACOG’s “early onset in life” criterion), nor is it characterized by disease traits that map to the other ACOG severity criteria. Similarly, Bloom syndrome met the “detrimental effect on quality of life” criterion because it is associated with high rates of cancer and immune deficiency leading to recurrent infections, but not with any of the other traits defining ACOG’s severity criteria. Given ACOG’s recommendation for panethnic cystic fibrosis carrier screening (11) and for Bloom syndrome carrier screening in Ashkenazi Jewish individuals (13), it appears that ACOG supports carrier screening for conditions that meet at least one of its four severity criteria.

Initial GC and MD discordance in ascribing disease traits to genes explains why five conditions did not meet any ACOG criteria despite being categorized as severe or moderate. In order for a trait to be mapped to ACOG criteria, all GCs and MDs assessing each gene had to initially agree to the trait. However, the phenotypic spectrum caused by variable expressivity was a common cause of initial discordant classification. For example, 21-hydroxylase congenital adrenal hyperplasia (21-OH CAH) can present in a severe form characterized by life threatening salt-wasting crises, or a less severe but more prevalent form characterized by physical malformation (ambiguous external genitalia). This variability in presentation led some GCs and MDs to initially assign disease traits differently for 21-OH CAH, and although consensus severity categorization was reached, initial discordant traits precluded mapping to ACOG criteria and resulted in 21-OH CAH not meeting any ACOG criteria. The severity algorithm applied here takes variable expressivity into consideration and focuses on traits present in at least 25% of individuals with mutations in a given gene, but the complexity of phenotypic presentation in many rare diseases exemplifies one difficulty in interpreting ACOG criteria.

### Utility of Screening for Moderate Severity Conditions by ECS

The term “moderate” as a descriptor of severity may connote to some that the disease does not rise to a level of significance for a couple wanting to know their risk for having a pregnancy affected by genetic disease. However, we found that 94.4% of moderate severity conditions met at least one of ACOG’s four severity criteria. As an example, *GJB2*, variants in which cause non-syndromic hearing loss and deafness, was categorized as having moderate severity. Early identification and interventions for individuals with hearing loss are critical; hearing loss complicates childhood development in language, socialization, academic performance, and most importantly human development and self-actualization (14-17). Interventions such as cochlear implants can effectively avert these developmental delays (17). Previous studies have also shown that couples find value in screening for moderate severity conditions. A majority of couples identified by ECS to be at risk for pregnancies affected by moderate severity conditions made reproductive and pregnancy management decisions for future family planning based on knowledge of such risk (6, 7). In addition, patients preferred to receive ECS results on conditions in all severity categories, including moderate, reporting that severity classifications informed their choice (5).

### Complexity of ECS panel design

ECS panel design is a complicated endeavor that must take into account a number of factors. (18-21). ACOG suggests several criteria other than those related to severity that should guide ECS panel design, including a carrier frequency of one in 100 or greater, a well-defined phenotype, and the ability to diagnose the disease prenatally (2). Clear definitions of ACOG criteria are needed to ensure consistency of ACOG criteria interpretations across different laboratories. In addition to the in-depth analysis of the severity criteria provided by this study, other criteria have been systematically evaluated. Ben-Shachar et al. (2019) and Guo and Gregg (2019) concluded that a panel meeting the “carrier frequency of 1 in 100 or greater” criterion would include approximately 40 conditions when that threshold is applied to any ethnicity (19, 22). Balzotti et al (2020) examined the “well-defined phenotype” criterion by applying the ClinGen framework for gene-disease association evidence (23) to more than 200 genes commonly included on commercial ECS panels, and found that the vast majority had the highest level gene-disease association evidence (24). These studies, taken together with ours, provide evidence-based interpretations of ACOG panel design criteria and act as guidance for laboratories in the design of ECS offerings.

## Limitations

This study has limitations that should be noted. First, the GC and MD participants were selected to review the conditions because of their expertise in pediatrics and medical genetics, but their application of the severity algorithm may not replicate application by other clinicians with similar or different expertise. Second, the MDs were not blinded to the final GC classifications, so it is possible that their classification was influenced by that of the GC pair. However, this paradigm imitates clinical practice, in which GCs (or other providers) routinely present initial findings to their clinical colleagues as the entire care team manages the patient. Third, genes were assessed based on the available literature, which may skew toward more severe disease presentations, particularly in the case of rare diseases. As understanding of phenotypes evolves, severity classifications may change. And fourth, the mapping of disease traits to ACOG criteria was not exhaustive, rather, it was limited to the traits in the algorithm. However, these traits were themselves determined by genetics professionals with experience in rare disease (4).

## Conclusions

This study applied a systematic and transparent process to evaluate the severity of genes on a commercially-available ECS panel by engaging with genetics providers with expertise in rare disease. The severity algorithm applied in this study brings clarity to ACOG’s severity criteria and can be used consistently across laboratories when considering genes for inclusion on ECS panels.

## Data Availability

The datasets generated during and/or analyzed during the current study are available from the corresponding author on request.

## Acknowledgements

The authors thank Danielle Fanslow and Anna Gardiner for design and editorial assistance. This study was funded by Myriad Women’s Health.

## Notes

### Competing Interest Statement

Aishwarya Arjunan, Holly Bellerose, Katherine Johansen Taber, Raul Torres, Jennifer Tarpinian, and Gabriel A. Lazarin are all current or former employees of Myriad Women’s Health. 
Jodi D. Hoffman, Brad Angle, Robert Nathan Slotnick, Brittany N. Simpson, Andrea M. Lewis, Pilar L. Magoulas, Kelly Bontempo, Jeanine Schulze, Jennifer Tarpinian, Jessica A. Bucher, Richard Dineen, and Allison Goetsch all received honoraria from Myriad Women’s Health to participate in this study. 
There are no other conflicts of interest to declare. 

### Funding Statement

No external funding was received.

